# Comparative Effects of E-cigarette Aerosol on Periodontium

**DOI:** 10.1101/2021.01.21.21250255

**Authors:** Fangxi Xu, Eman Aboseria, Malvin N Janal, Smruti Pushalkar, Maria V Bederoff, Rebeca Vasconcelos, Sakshi Sapru, Bidisha Paul, Erica Queiroz, Shreya Makwana, Julia Solarewicz, Yuqi Guo, Deanna Aguallo, Claudia Gomez, Donna Shelly, Yindalon Aphinyanaphongs, Terry Gordon, Patricia Corby, Angela R. Kamer, Xin Li, Deepak Saxena

**Affiliations:** Department of Molecular Pathobiology, New York University College of Dentistry, New York, NY 10010, USA; Department of Epidemiology and Health Promotion, New York University College of Dentistry, New York, NY 10010, USA; Department of Periodontology and Implant Dentistry, New York University College of Dentistry, New York, NY 10010, USA; Department of Population Health, New York University School of Medicine, New York, NY 10016, USA; Department of Medicine, New York University School of Medicine, New York, NY 10016, USA; Department of Environmental Medicine, New York University School of Medicine, New York, NY 10016, USA; Department of Oral Medicine, University of Pennsylvania, School of Dental Medicine, Philadelphia, PA 19104, USA

**Keywords:** E-cigarettes, aerosol, smoking, microbiome, periodontal disease, host response

## Abstract

**Introduction:** Tobacco use is one of the main causes of periodontitis. E-cigarettes are gaining in popularity, and studies are needed to better understand the impact of e-cigarettes on oral health. Objective: To perform a longitudinal study to evaluate the adverse effects of e-cigarettes on periodontal health.

**Methods:** Naïve e-cigarette users, cigarette smokers, and non-smokers were recruited using newspaper and social media. Demographics, age, gender, and ethnicity, were recorded. Participants were scheduled for two visits 6 months apart. At each visit, we collected data on the daily frequency puffs of an e-cigarette, the number of cigarettes smokes, and other parameters, such as alcohol consumption. Carbon monoxide levels, cotinine levels, salivary flow rate, probing depth, and bleeding on probing were determined at both baseline and follow-up visits. P-values less than 0.05 were considered statistically significant.

**Results:** We screened 159 subjects and recruited 140 subjects. One-hundred-one subjects (31 cigarette smokers, 32 e-cigarette smokers, and 38 non-smokers) completed every assessment in both visits. The retention and compliance rate of subjects was 84.1%. The use of social media and craigslist was significant in recruiting e-cigarette subjects. Ethnicity and race differed between cohorts, as did average age in the male subjects. Carbon monoxide and salivary cotinine levels were highest among cigarette smokers. Bleeding on probing and average probing depths similarly increased over time in all three cohorts. Increase in the rates of severe periodontal disease were significantly higher in cigarette smokers and e-cigarette users than non-smokers. Confounding factors were subjects’ age as most of the e-cigarette group were much younger than cigarette smokers.

**Conclusion:** Among the recruited participants, periodontal severity status after 6 month was significantly worse in cigarette smokers and e-cigarette smokers than non-smokers. This study design and protocol will assist in future larger studies on e-cigarette and oral health.

## Introduction

Periodontitis, also known as gum disease, is a chronic, polymicrobial inflammatory disease affecting the tissue supporting the tooth. One of the main risks for periodontitis is smoking, as it alters the microbiome (Eggert et al. 2001) of the oral cavity and the host immune response (Lee et al. 2012), causing the oral tissue to become vulnerable and susceptible to disease. Previous studies have demonstrated that the use of tobacco-containing products could potentially lead to oral manifestations, such as mucosal lesions (e.g., leukoplakia, candidiasis, nicotine stomatitis), plaque formation, teeth staining, gingivitis, periodontitis, tooth loss, failure of prosthetic and surgical treatments, and increased risk of oral cancer (Chaffee 2019; Couch et al. 2016; Johnson and Bain 2000). Over the years, conventional cigarette smoking has declined; however, the use of emerging tobacco products, such as electronic cigarettes (e-cigarettes) has increased (Centers for Disease Control and Prevention 2016). E-cigarettes are non-combustible battery-operated devices that allow users to inhale an aerosol mixture that typically contains propylene glycol and/or glycerin with or without nicotine and other additives (Breland et al. 2017). It has been proposed that e-cigarettes serve as a strategy of smoking cessation or a less harmful replacement for conventional cigarettes. However, the data is inconclusive (Charlotte Wells 2017). Switching from smoking to e-cigs reduces the number of cigarettes smoked; however, it does not result in complete withdrawal, and the risk of developing smoking-related diseases, particularly oral diseases, remains a high possibility (Malas et al. 2016; Tomar et al. 2015). Moreover, the CDC recently reported 2668 hospitalized e-cigarettes use-associated lung injury cases or deaths (Centers for Disease Control and Prevention 2020, February 25). Among those cases, 15% of patients were under 18 years old, and 37% of patients were 18 to 24 years old (Centers for Disease Control and Prevention 2020, February 25). These pathologies suggest that e-cigarettes can significantly damage various tissues, including oral tissues. As the popularity of e-cigarettes use increases, and the potential for damage exists, it is necessary to investigate the impact of e-cigarette use on oral health.

The e-cigarette aerosol includes, but is not limited to, tobacco-specific nitrosamines, aldehydes, metals, and volatile organic compounds (Cheng 2014). These compounds could potentially alter the oral microbiome and have adverse effects on oral health. Disturbance of the oral microbiome, particularly commensal microorganisms, might lead to dysbiosis and increase pathobionts, which might lead to oral diseases, such as periodontal disease. Dysbiosis might, in turn, activate different inflammatory pathways and, subsequently, lead to systemic health conditions, such as respiratory (Chun et al. 2017; Clapp and Jaspers 2017), immune (Reidel et al. 2018), and cardiovascular complications (Benowitz and Fraiman 2017). Furthermore, our recent study showed that e-cigarette aerosol exposure caused elevated concentrations of proinflammatory cytokines (IL)-6 and IL-1β, thus potentially increasing susceptibility to periodontal disease (Pushalkar et al. 2020).

Clinical parameters of periodontal inflammation include clinical attachment loss, increased probing depth (PD), and bleeding on probing (BoP) (Jeong et al. 2020; Lang and Bartold 2018). Studies have shown that clinical parameters of periodontitis are poorer in cigarette smokers compared to non-smokers (Al-Wahadni and Linden 2003). However, there is limited information on the impact of e-cigarette use on oral health, particularly periodontal status. The current longitudinal study is designed to present a demographic description of our population and compare the clinical periodontal status among cigarette smokers, e-cigarette users, and non-smokers. The primary hypothesis is that clinical parameters of periodontal disease are worse in cigarette smokers and e-cigarette users compared to non-smokers. The findings of this study will help to understand the potential risks associated with e-cigarette use.

## 2. METHODS

### 2.1. Ethical guidelines

The approval of the study protocol, informed consent form(s), and all subject materials were obtained by the Institutional Review Board (IRB) of the New York University Langone Medical Center. Before any study-related assessment, the participants received a detailed explanation of the research study and procedures. The informed consent of each participant was obtained prior to sample collection, and a copy of the consent form was provided to each participant for their record. Information regarding the risks and possible benefits of study participation was provided, and participants were informed that they might withdraw consent at any time throughout the course of the study. All STROBE guidelines were followed.

### 2.2. Study design and participants

The present study aimed to compare clinical indicators (PD, BOP, CAL) of periodontitis among cigarette smokers, e-cigarette users, and non-smokers. Study visits were conducted at the NYUCD Bluestone Center for Clinical Study. Upon obtaining informed consent and completion of a standardized oral health questionnaire, further social, medical, and dental history was recorded. Subjects were asked to report on the frequency and intensity of tobacco and alcohol use, previous and current health conditions, surgeries, medications, and symptoms of existing conditions. Carbon monoxide levels were assessed, and saliva was collected for the determination of cotinine levels. Periodontal examinations were performed by a calibrated examiner, and subgingival plaque and saliva samples were collected for microbiome analysis (reported elsewhere) (Pushalkar et al. 2020). A follow-up visit (V2) was scheduled 6 months after the baseline visit (V1), and the protocol was repeated along with the assessment of adverse events. Participant charts were assigned an identification number and secured at the NYU’s Bluestone Center for Clinical Research.

To be eligible for the study, participants were required to meet conditions specific to each cohort. A cigarette smoker was defined as someone who, at the time of the study, smoked at least 10 cigarettes daily for a period of 12 months or more. E-cigarette users were defined as a non-cigarette smoker who used minimum of 0.5–1 e-cigarettes daily for minimum of the last 6 months. Lastly, a non-smoker was defined as someone who never smoked a cigarette or used an e-cigarette in their lifetime.

### 2.3. Participants enrollment, recruitment, and eligibility

Recruitment of participants was managed by the study coordinator and personnel from New York University’s Bluestone Center for Clinical Research. Study flyers were displayed at NYUCD Television screens, NYU primary care, and dental clinics, as well as the Health and Hospital Corporation’s primary care sites. Additionally, the study advertisement was posted in local newspapers on Craig’s list and Facebook, which has been an effective tool for recruitment. Participants were required to be 21 years of age, to have a minimum of 16 teeth, including eight posterior teeth, and diagnosed with mild, moderate, or severe periodontal disease (Jeong et al. 2020; Lang and Bartold 2018). The exclusion criteria were as follows: (a) a medical condition (including uncontrolled diabetes and HIV); (b) recent febrile illness that delays or precludes participation; (c) pregnancy or lactation; (d) history of radiation therapy to the head and neck region; (e) antibiotic use or professional dental cleaning within 1 month; (f) enrollment in other studies; (g) or presence of oral mucosal lesions, such as leukoplakia, herpes labialis, and candidiasis. In addition, non-smoker subjects were excluded from the study if the carbon monoxide (CO) level was at least 7 parts per million (ppm), calling into doubt their non-smoking status.

Among the 159 subjects who attended the screening visits, a total of 119 subjects participated in our study; 39 non-smokers, 40 exclusively conventional cigarette smokers, and 40 exclusively e-cigarette users successfully enrolled and completed all the assessments of baseline visits. Of these participants, 101 (38 non-smokers, 31 cigarette smokers, and 32 e-cigarette users) have completed the follow-up examination 6 months after the baseline visit (Fig. 1). Participants who did not complete the follow-up visit were either lost to follow-up, withdrew for personal reasons (such as relocation).

**Figure 1.**
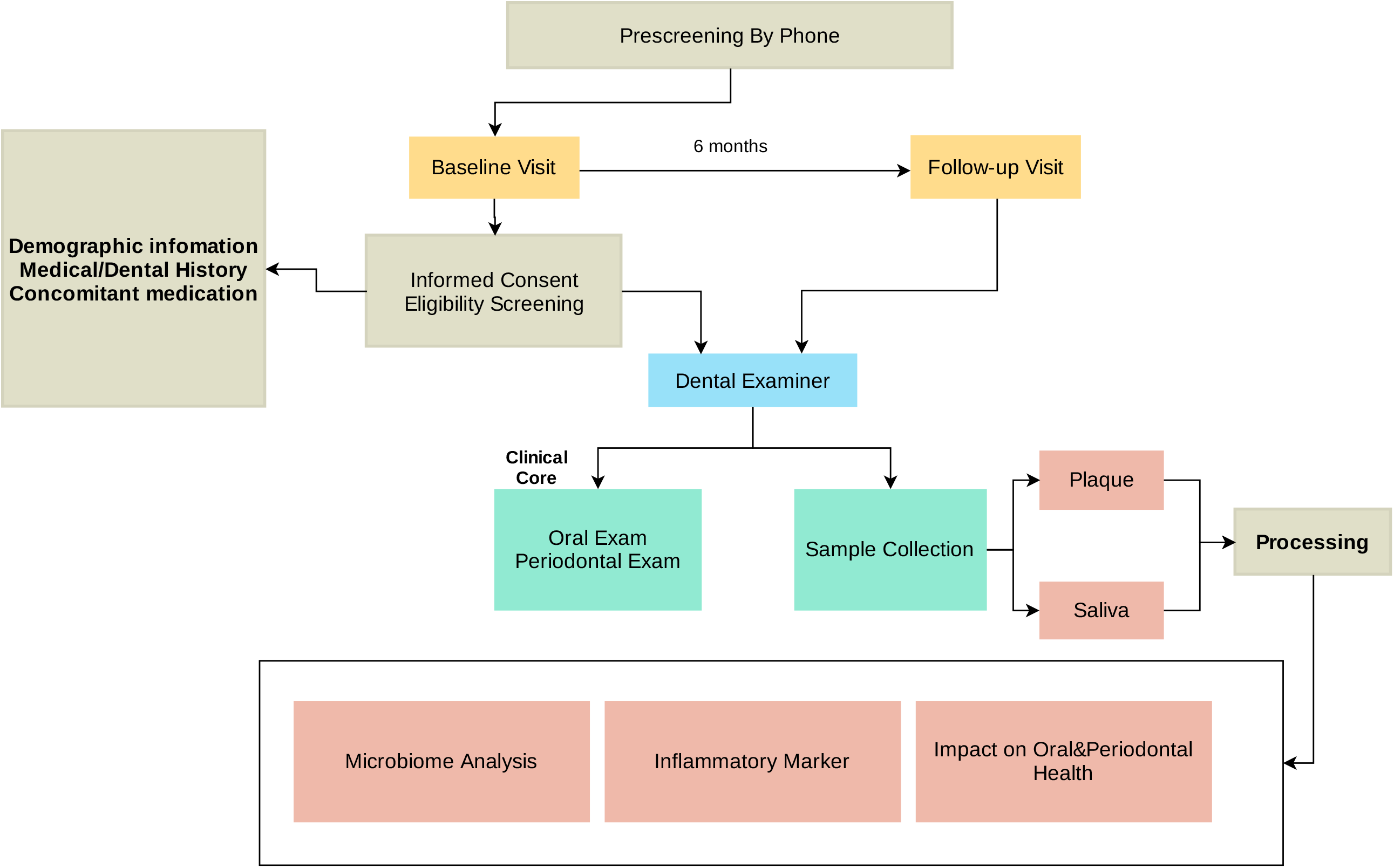
Schematic showing the flow of subject recruitment and sample collection. The types of samples collected and their transit from collection points to a processing laboratory where they were labeled, entered into an inventory database.

### 2.4. Questionnaire

A questionnaire was developed from the Center for Disease Control and Prevention (CDC) oral health questionnaire (Centers for Disease Control and Prevention 2011, July 22) and completed at baseline and follow-up visits. The questionnaire included eight questions related to periodontal health and past treatment, tooth status, and how many times they used floss and mouthwash during the preceding 7 days.

### 2.5. Clinical data collection

Subject’s sex, age, ethnicity, race, nicotine (conventional cigarette and e-cigarette [daily puffs]), and alcohol use history were recorded. Subjects who were eligible and included in this study were asked to follow up daily via specially created text messaging applications to monitor smoking and e-cigarette status for compliance. The information collected was secured by the REDCap database and Twilio software. The subject’s identification was encrypted, and the information was transferred to the NYUCD database. The database includes the medical history, dental history, and periodontal status of the subjects.

### 2.6. Assessment of CO levels

To confirm the smoking status of each participant, carbon monoxide (CO) levels were tested by CO Smokerlyzer (Smokerlyzer, Covita, Santa Barbara, CA) according to the manufacturer’s instructions. Participants were instructed to inhale deeply and hold their breath for fifteen seconds before slowly exhaling into the device. Based on the CO test results, participants were categorized into one of five groups: non-smoking (NS) (0–6 ppm), low addicted smokers (LAS) (10–15 ppm), moderately addicted smokers (MAS) (16–25 ppm), heavily addicted smokers (HAS) (26–35), and very heavily addicted smokers (VHAS) (≥ 36 ppm) (Pushalkar et al. 2020).

### 2.7. Oral examination

Oral examination was performed by three different calibrated periodontists or dental hygienists. Oral examination was completed at each visit and included: mucosal assessment of lower and upper lip, hard and soft palate, uvula, the floor of the mouth, tongue, tonsils, and labial and buccal mucosa. If any abnormality (such as candidiasis, herpes labialis, aphthous stomatitis) was present, the participant was referred to an oral medicine specialist.

### 2.8. Gingival and periodontal assessment

A full mouth examination was performed to assess the periodontal condition. Periodontal measurements were recorded at six sites per tooth (mesio-buccal, buccal, disto-buccal, mesio-lingual, lingual, and disto-lingual) on all teeth present and included the following: (1) probing depth (PD) defined as the distance from the free gingival margin to the depth of the pocket; (2) distance from the free gingival margin to the cement enamel junction (CEJ); and (3) presence or absence of bleeding on probing (BOP). Clinical attachment loss (CAL) was then calculated by subtracting the CEJ measurement from the PD. For analysis, the percentage of bleeding sites was determined by dividing the number of sites that bled by the total number of sites sampled and multiplying by 100. Probing depth and CAL were summarized as the average PD and CAL among the sampled sites.

The classification of mild, moderate, or severe periodontal disease followed the definition given by the CDC in collaboration with the American Academy of Periodontology (CDC-AAP) (Eke et al. 2013). Mild periodontitis was defined as ≥ two interproximal sites with ≥ 3 mm attachment loss, and ≥ 2 mm interproximal sites with probing depth ≥ 4 mm (not on the same tooth), or one interproximal site with PD ≥ 5 mm. Moderate periodontitis was defined as two or more interproximal sites with ≥ 4 mm clinical AL (not on the same tooth), or two or more interproximal sites with PD ≥ 5 mm, also not on the same tooth. Severe periodontitis was defined as having two or more interproximal sites with ≥ 6 mm AL (not on the same tooth), and one or more interproximal site(s) with ≥ 5 mm PD [20].

### 2.9. Saliva sample collection and flow rate assessment

Participants were asked to chew paraffin wax pellets (Gleegum, Verve Inc., Providence, RI) to stimulate salivary secretion. After chewing gum for 30 s to 1-min, participants were asked to expectorate 10 ml saliva into a sterile graduated 50 mL centrifuge tube on ice. The amount of saliva was measured after 5 min. If the measured amount was less than 5 mL, participants were asked to keep expectorating. The salivary flow rate was calculated based on recorded data at 5 min. Saliva samples were stored on ice and delivered to the clinical site’s laboratory for processing. Some saliva (1 mL) was utilized immediately for the cotinine level evaluation (Nic Alert kit, Salimetrics, State College, PA). Retained samples were aliquoted, preserved with phenylmethylsulfonyl fluoride (PMFS), and subjected to aprotinin immune mediator analysis. Aliquots were also saved for microbiome analysis. All the samples were stored at -80°C.

### 2.10. Plaque sample collection

Subgingival plaque samples were collected before periodontal probing by the study clinician using the stroke technique with sterile Gracey mini-curette from the distal and mesial aspects of eight posterior teeth. The samples were then placed into individual sterile 2 mL microcentrifuge tubes with transport (TE) buffer and kept on ice until delivered to the lab, where PMFS buffer and aprotinin were added, labeled, and stored at -80°C.

### 2.11. Statistical analysis

All data were exported from NYULMC REDCap, and statistical analysis was performed using IBM SPSS (v26, IBM Corp., Armonk, NY). Analysis of continuous measures (a measure of CO, salivary flow rate, PD, BOP, and CAL) compared means from the three cohorts over time using a two-way mixed model analysis of variance (ANOVA) followed by Tukey’s honestly significant difference (HSD) test. If confronted with heterogeneous variances, the Kruskal-Wallis or Welch test was substituted. Differences between groups in rates of periodontal diagnosis were evaluated using the chi-square test, and changes in diagnosis over time within groups were evaluated using the McNemar test. Logistic regression was then used to evaluate confounding between-group differences in clinical and demographic variables. It was estimated that a sample size of 40 per cohort was sufficient to detect a one standard deviation difference in a two-tailed independent samples t-test with a power of 99%. P-values less than 0.05 were considered statistically significant.

## RESULTS

### Participant’s demographics

A total of 101 subjects completed the baseline and 6-month follow-up evaluations and sample collections: 31 were cigarette smokers, 32 were e-cigarette smokers, and 38 were non-smokers. The demographic characteristics of the study subjects are shown in Table 1. Seventy percent of the subjects were male. Among males, non-smokers were significantly younger than e-cigarette smokers, and e-cigarette smokers were significantly younger than cigarette smokers. Most non-smokers were Asian, most cigarette smokers were Black, and most e-cigarette smokers were White.

**Table 1.** Demographics

|  | **Cigarette smokers** | **E-cigarette smokers** | **Non-Smokers** | **p value** |
| --- | --- | --- | --- | --- |
| **N** | 31 | 32 | 38 |  |
| **Sex (% male)** | 77.4 | 78.1 | 57.9 | 0.11 |
| **Age (yr), M (SD)** |  |  |  |  |
| *Male* | 46.9 (10.1)<sup>b</sup> | 36.0 (9.8)<sup>c</sup> | 28.8 (6.1)<sup>a</sup> | <0.001* |
| *Female* | 46.6 (11.5) | 38.7 (10.6) | 39.0 (14.1) | 0.40 |
| **Ethnicity (% Hispanic)** | 6.5 | 22.6 | 23.7 | 0.13 |
| **Race (%)** |  |  |  | 0.002 |
| *White* | 32.3 | 56.3 | 26.3 |  |
| *Black* | 54.8 | 34.4 | 28.9 |  |
| *Asian* | 9.7 | 6.3 | 42.1 |  |
| *Other* | 3.2 | 3.1 | 2.6 |  |

\* One-way analysis of variance and Tukey post-hoc test.
<sup>a;b;c</sup> Unlike superscripts indicate significantly different group means( $p<0.05$ )

### Tobacco and alcohol use

By design, each cohort was enrolled considering the inclusion criteria for the smoking behavior. Table 2a shows that e-cigarette smokers consumed an average of less than one cartridge per day at each study visit. However, the average puffs per day of e-cigarettes declined between the baseline visit and follow-up visit (p = 0.03). By contrast, cigarette smokers maintained a constant average use over time, of about 13 cigarettes per day (p = 0.70).

**Table 2a.** Smoking Behavior

|  | **Baseline Visit** | **Follow-up Visit** | **p value** |
| --- | --- | --- | --- |
| **E-cigarette smoker** |  |  |  |
| *E-cigarettes/day* | 0.78 (0.25) | 0.74 (0.51) | 0.70 |
| *Puffs/use* | 151.3 (104.4) | 94.4 (99.1) | 0.03 |
| **Cigarette smoker** |  |  |  |
| *Cigarettes/Day* | 13.5 (4.8) | 12.3 (4.4) | 0.17 |

Table 2b shows that approximately half of the subjects in each study cohort reported using alcohol on both visits. Each cohort reported drinking about two times per week, consuming two or three drinks each time. Although not reaching statistical significance, e-cigarette smokers tended to drink more often than others (p = 0.1).

**Table 2b.** Alcohol Consumption

|  | *Alcohol User at f/u,*<br>*n(%)* | *Drinking (days/week),*<br>*Mean(SD)\** | *# Drinks/drinking*<br>*day, Mean(SD)\** |
| --- | --- | --- | --- |
| **Cigarette smokers** | 14 (48.4) | 2.0 (1.2) | 2.8 (2.3) |
| **E-cigarette smokers** | 15 (53.1) | 2.4 (1.2) | 4.1 (2.3) |
| **Non-Smokers** | 18 (50.0) | 1.5 (1.2) | 2.6 (2.3) |
| **p value** | 0.93 | 0.10 | 0.16 |

*\*The analysis of alcohol use (days/week and # drinks/day) is based on the consuming participants.*

### The CO, cotinine level, and saliva flow rate across groups

Cigarette smoking subjects showed higher mean levels of carbon monoxide than e-cigarette smokers or non-smokers (Fig. 2A, 20.8 vs. 5.8 and 2.8 ppm, respectively, p < 0.001), as well as higher mean levels of salivary flow rate (Fig. 2B, 3.1 vs. 2.2 and 2.5 mL/min, respectively, p = 0.02). Consistent with their smoking behaviors, salivary cotinine levels were higher in cigarette smokers than e-cigarette smokers, who were higher than non-smokers at each test period (Fig. 2C Kruskal-Wallis test, all p < 0.001). Figure 2C also shows a reduction in cotinine levels over time in the cigarette smokers (paired sample t-tests p = 0.002). Analysis failed to show an interaction between group and time on any of these measures.

**Figure 2.**
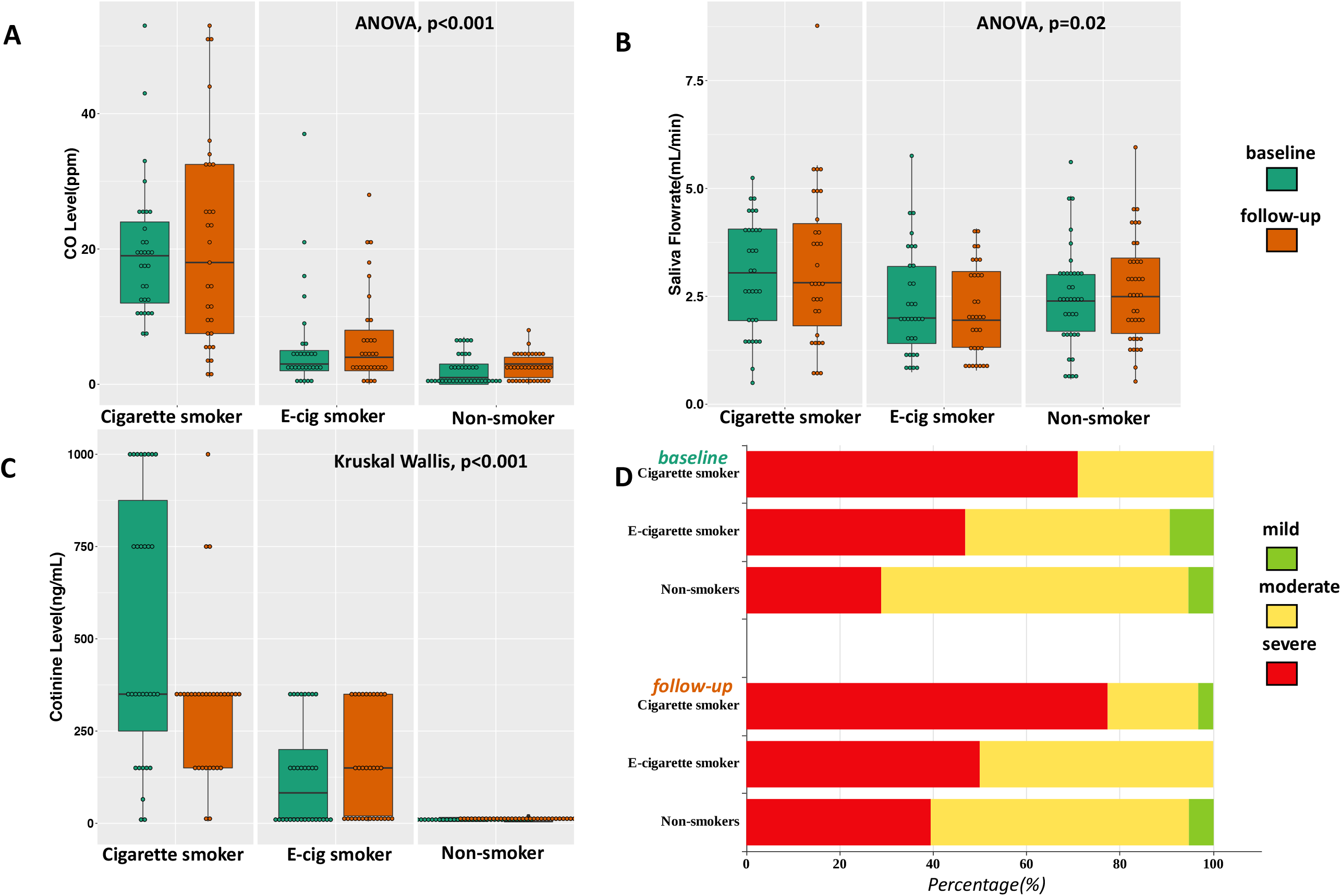
(A) Levels of breath carbon monoxide (ppm) across the subjects in the cigarette smokers, e-cigarette smokers, and the Non-smokers at base line and 6 month follow-up: 0-6 ppm, Borderline (BdL): 7-9 ppm, low addicted smoker (LAS): 10-15 ppm, moderate addicted smoker (MAS): 16-25 ppm, heavily addicted smoker (HAS): 26-35 ppm and very heavily addicted smoker (VHAS): 36+ ppm. (B) Saliva flow rate in three cohort at base line and follow-up. (C) Distribution of salivary cotinine levels in the participants of the three cohorts. (D) Prevalence of periodontal disease in all three cohorts from baseline visits to follow-up visits.

### Bleeding on probing, pocket depth, clinical attachment loss, and periodontal disease status

The mean percentage of bleeding on probing (BoP) in the three cohorts was increased similarly over time from 56% at baseline to 64% at follow-up (Table 3, p = 0.03). The mean probing depth and clinical attachment loss (CAL) were significantly greater in the cigarette smoker group than the non-smoker group or e-cigarette smoker group at both visits (Table 3). While there was an increase in probing depth over time from 2.9 to 3.1 mm, a change that was comparable for the three cohorts, CAL was stable over time. As age, sex, and race were not evenly distributed among the cohorts, we evaluated confounding variables between cohort and demographic effects. Age was not related to either bleeding on probing or probing depth, and so analyses of cohort differences in probing depth were unchanged when adjusted for age and sex. These results suggest similar levels of gingival health and changes in gingival health overtime in each cohort, but more attachment loss in cigarette smokers than other cohorts. Periodontal disease diagnosis also varied among the cohorts studied. Table 3 shows that the prevalence of severe periodontal disease was higher among cigarette smokers than either other cohorts, at both visits (baseline, p = 0.002; follow-up, p = 0.006). The prevalence of severe periodontal disease increased in all three cohorts from baseline visits to follow-up visits (Fig. 2D), multiple comparison showed no significant changes (McNemar test: Cigarette smoker, p = 0.69; e-cigarette smoker, p = 1.00; Non-smoker, p = 0.22) however all the mild periodontal subject progressed to moderate to severe periodontitis in 6 month follow-up analysis. Prevalence of severe periodontal disease also increased with age (baseline r = 0.43; follow-up r = 0.47, both p < 0.001) (Table 3b). These results suggest that e-cigarettes smokers has higher risk of periodontal disease progression as compared to non-smokers.

**Table 3a.** Rates of BoP and periodontal disease severity, and levels of, probing depth and clinical attachment loss as a function of cohort and time

|  | Visit | Non-Smokers | E-cigarette smokers | Cigarette smokers | p value |
| --- | --- | --- | --- | --- | --- |
| *Bleeding on Probing (%)*, Mean(SD)* | Baseline | 52.6(32.1) | 53.0 (32.5) | 61.5 (27.6) | 0.03 |
|  | Follow-up | 68.3(24.4) | 57.0 (32.6) | 66.2 (30.5) |  |
| *Probing depth (mm)*, Mean (SD) | Baseline | 2.7 (0.4)<sup>a</sup> | 3.0 (0.6)<sup>a</sup> | 3.1 (0.7)<sup>b</sup> | 0.001* |
|  | Follow-up | 2.9 (0.4)<sup>a</sup> | 3.1 (0.7)<sup>a</sup> | 3.2 (0.6)<sup>b</sup> | 0.01* |
| *Clinical attachment loss (mm)*, Mean (SD) | Baseline | 2.2 (0.9)<sup>a</sup> | 2.8 (1.5)<sup>a</sup> | 3.5 (1.1)<sup>b</sup> | <0.001* |
|  | Follow-up | 2.2 (0.7)<sup>a</sup> | 3.1 (1.4)<sup>a</sup> | 3.4 (1.1)<sup>b</sup> | <0.001* |
| *Periodontal disease (% progressed)* + | Follow-up | 18 | 29 | 44 | NA |

\* Welch test for heterogeneous variances and Tukey post-hoc test.
<sup>a;b;c</sup> Unlike superscripts indicate significantly different group means( $p<0.05$ ).
+Cases with mild and moderate periodontal disease used in this analysis.

**Table 3b.** Logistic regression analysis relating rates of severe periodontal disease to cigarette smoking status and age.

| Baseline Visit |  |  |  | Follow-up Visit |  |  |  |
| --- | --- | --- | --- | --- | --- | --- | --- |
| *Step1\** |  | OR | p value | *Step1\** |  | OR | p value |
|  | Cigarette smoking | 6.0 | <0.001 |  | Cigarette smoking | 5.3 | 0.002 |
| *Step2\** |  |  |  | *Step2\** |  |  |  |
|  | Cigarette smoking | 2.8 | 0.08 |  | Cigarette smoking | 1.9 | 0.31 |
|  | Age(y) | 1.07 | 0.003 |  | Age(y) | 1.09 | <0.001 |

*\*Smoking status was entered into the regression on step 1, and age was then added in step 2. When adjusting age, smoking status no longer predicts periodontal severe periodontal disease.*

## DISCUSSION

Although a great deal of literature is available on conventional tobacco products, limited data is available on the effects of e-cigarette on oral health. To address this, we conducted a clinical study to determine the impact of e-cigarette use on oral health, particularly periodontal health. The initial interaction of e-cigarette aerosol mixtures occurs largely in the oral cavity, where nicotine and other compounds are expected to be most active, and the exposure is most intense. A recent online survey of 543 e-cigarette users indicated that most negative health effects were observed in the mouth and throat (Hua et al. 2013). We reported previously that the periodontal inflammatory indicators were elevated in cigarette smokers relative to individuals who use e-cigarettes and to non-smokers (Pushalkar et al. 2020).

In our clinical study for recruitment we used electronic social media and we were very successful in recruiting naive e-cigarette subjects. One hundred fifty nine subjects attended the screening visits and 120 were recruited, 101 completed the follow-up assessments after 6 months. Among this 24 failed the initial screening, and 15% withdrew or were lost to the follow-up visit. In the analysis, we only included subjects who completed both visits.

The cigarettes smoking behavior did not change in the use of cigarette or e-cigarette per day but there was a significant change in the number of puffs per e-cigarette (151.3 puffs at baseline and 94.4 puffs at 6 month follow up), suggesting more intensive puffing and higher consumption of nicotine per puff as they get adapted to e-cigarette. Studies have shown that puffing patterns associated with nicotine strength or e-liquids and voltage used in e-cigarette which result in higher toxicant exposure (Cox et al. 2016; Dawkins et al. 2016; Farsalinos et al. 2018).

The prevalence of severe periodontal disease was higher than expected in all cohorts (Fig 2D). This is likely due to the inclusion criterion that required at least mild levels of periodontal disease. Nevertheless, the rate of severe periodontal disease increased in all three groups from baseline visit to follow-up visit, however, this more severe in cigarette smokers and e-cigarettes users. When comparing bleeding on probing did between the e-cigarette group and non-smokers, no significant difference was detected; however, probing depth and clinical attachment loss was much higher in the e-cigarette group. Overall, the percentage of severe periodontal disease was much higher in the e-cigarette group than among the non-smokers (Table 3a).

A limitation of the study design was that the groups were not matched for age; although it is difficult as e-cigarette users are much younger than the cigarette smokers but this should be a considered in all future e-cigarette clinical research. E-cigarette users were much younger than the cigarette smokers, and that cofounded some of our findings (Garnett et al. 2020; Jeong et al. 2020; Omoike and Johnson 2020; Subica et al. 2020; Vallone et al. 2020; Walker et al. 2020; Wamamili et al. 2020). Age was also considered a confounder of the relationship between cigarette smoking status and the rate of severe periodontitis. As such, age appears to be the more parsimonious explanation of higher rates in those participants. Another variable that could have led to differential outcomes is the ethnic group, which was not evenly distributed in the study cohorts. We notice that most non-smokers were Asian, most cigarette smokers were black, and most e-cigarette smokers were white, suggesting disparity among smokers and e-cigarette users. It has been reported by the CDC that US Blacks and Hispanics show poorer oral health compared to Whites and Asians (Centers for Disease Control and Prevention 2016, May 17). Nevertheless, while the majority of the subjects in the non-smoker group were Asian, e-cigarette users were primarily White, and cigarette smokers were primarily Black, the analysis showed effects of race on periodontal status.

Modified questionnaires, with more precise information on the subject’s social practices, including alcohol usage and their dental hygiene routine, will control for confounding factors in the study. The study design could be further improved if social and education status were evaluated to check whether there is a relationship between higher education, better oral health, and e-cigarette use.

To our knowledge, this is the first clinical research report on the oral health impacts of vaping (e-cigarette use) relative to cigarette smokers and non-smokers. The described study design and its limitations can guide future larger studies on e-cigarette use.

## Data Availability

All data will be available as per NIH guideline upon publication

## ACKNOWLEDGEMENTS

This research project was supported by NIH grants DE025992 (DS, XL), DE027074 (DS, XL), CA206105 (DS) and the NYU Mega grant initiative (DS, XL).

## AUTHOR CONTRIBUTIONS

FX and EA: carried out REDCap data entry, data analyses and interpretation, manuscript preparation; S.P and BP: carried out sample collection, data analyses and interpretation, manuscript preparation; MB, SS, SM, JS, YQ: carried out clinical data analysis, data entry, technical lab work and statistical analyses, manuscript preparation; M.L. statistical data analyses and critical review; EQ, RV: carried out subject recruitment; D.A. performed oral exam and clinical sample collection; C.G. performed oral exam and clinical sample collection; A.K. performed oral exam, clinical sample collection and analyses; D.S. performed subject recruitment; Y.A. managed REDCap, clinical data and electronic messaging system; T.G. assisted in aerosol generating machine and manuscript preparation; P.C. provided assistance in subject recruitment and clinical sample collection; X.L. and D.S. conceived, designed, supervised, analyzed, interpreted the study, provided critical review and manuscript preparation.

## References

Al-Wahadni A, Linden GJ. 2003. The effects of cigarette smoking on the periodontal condition of young jordanian adults. J Clin Periodontol. 30(2):132–137.

Benowitz NL, Fraiman JB. 2017. Cardiovascular effects of electronic cigarettes. Nat Rev Cardiol. 14(8):447–456.

Breland A, Soule E, Lopez A, Ramoa C, El-Hellani A, Eissenberg T. 2017. Electronic cigarettes: What are they and what do they do? Ann N Y Acad Sci. 1394(1):5–30.

Oral health questionnaire. 2011, July 22. [accessed]. https://www.cdc.gov/nchs/data/nhanes/nhanes_11_12/ohq.pdf.

Centers for Disease Control and Prevention. 2016. Chapter 1 introduction, conclusions, and historical background relative to e-cigarettes. Disparities in oral health. 2016, May 17. [accessed]. https://www.cdc.gov/oralhealth/oral_health_disparities/index.htm.

Outbreak of lung injury associated with e-cigarette use, or vaping. 2020, February 25. [accessed]. https://www.cdc.gov/tobacco/basic_information/e-cigarettes/severe-lung-disease.html#latest-information.

Chaffee BW. 2019. Electronic cigarettes: Trends, health effects and advising patients amid uncertainty. J Calif Dent Assoc. 47(2):85–92.

Charlotte Wells KF. 2017. Electronic cigarettes for the reduction or cessation of smoking: Clinical utility, safety, and guidelines. Ottawa: CADTH.

Cheng T. 2014. Chemical evaluation of electronic cigarettes. Tob Control. 23 Suppl 2:ii11–17.

Chun LF, Moazed F, Calfee CS, Matthay MA, Gotts JE. 2017. Pulmonary toxicity of e-cigarettes. Am J Physiol Lung Cell Mol Physiol. 313(2):L193–L206.

Clapp PW, Jaspers I. 2017. Electronic cigarettes: Their constituents and potential links to asthma. Curr Allergy Asthma Rep. 17(11):79.

Couch ET, Chaffee BW, Gansky SA, Walsh MM. 2016. The changing tobacco landscape: What dental professionals need to know. J Am Dent Assoc. 147(7):561–569.

Cox S, Kosmider L, McRobbie H, Goniewicz M, Kimber C, Doig M, Dawkins L. 2016. E-cigarette puffing patterns associated with high and low nicotine e-liquid strength: Effects on toxicant and carcinogen exposure. BMC Public Health. 16:999.

Dawkins LE, Kimber CF, Doig M, Feyerabend C, Corcoran O. 2016. Self-titration by experienced e-cigarette users: Blood nicotine delivery and subjective effects. Psychopharmacology (Berl). 233(15-16):2933–2941.

Eggert FM, McLeod MH, Flowerdew G. 2001. Effects of smoking and treatment status on periodontal bacteria: Evidence that smoking influences control of periodontal bacteria at the mucosal surface of the gingival crevice. J Periodontol. 72(9):1210–1220.

Eke PI, Dye BA, Wei L, Slade GD, Thornton-Evans GO, Beck JD, Taylor GW, Borgnakke WS, Page RC, Genco RJ. 2013. Self-reported measures for surveillance of periodontitis. J Dent Res. 92(11):1041–1047.

Farsalinos K, Poulas K, Voudris V. 2018. Changes in puffing topography and nicotine consumption depending on the power setting of electronic cigarettes. Nicotine Tob Res. 20(8):993–997.

Garnett C, Tombor I, Beard E, Jackson SE, West R, Brown J. 2020. Changes in smoker characteristics in england between 2008 and 2017. Addiction. 115(4):748–756.

Hua M, Alfi M, Talbot P. 2013. Health-related effects reported by electronic cigarette users in online forums. J Med Internet Res. 15(4):e59.

Jeong W, Choi DW, Kim YK, Lee HJ, Lee SA, Park EC, Jang SI. 2020. Associations of electronic and conventional cigarette use with periodontal disease in south korean adults. J Periodontol. 91(1):55–64.

Johnson NW, Bain CA. 2000. Tobacco and oral disease. Eu-working group on tobacco and oral health. Br Dent J. 189(4):200–206.

Lang NP, Bartold PM. 2018. Periodontal health. J Periodontol. 89 Suppl 1:S9–S16.

Lee J, Taneja V, Vassallo R. 2012. Cigarette smoking and inflammation: Cellular and molecular mechanisms. J Dent Res. 91(2):142–149.

Malas M, van der Tempel J, Schwartz R, Minichiello A, Lightfoot C, Noormohamed A, Andrews J, Zawertailo L, Ferrence R. 2016. Electronic cigarettes for smoking cessation: A systematic review. Nicotine Tob Res. 18(10):1926–1936.

Omoike OE, Johnson KR. 2020. Prevalence of vaping and behavioral associations of vaping among a community of college students in the united states. J Community Health.

Pushalkar S, Paul B, Li Q, Yang J, Vasconcelos R, Makwana S, Gonzalez JM, Shah S, Xie C, Janal MN et al. 2020. Electronic cigarette aerosol modulates the oral microbiome and increases risk of infection. iScience. 23(3):100884.

Reidel B, Radicioni G, Clapp PW, Ford AA, Abdelwahab S, Rebuli ME, Haridass P, Alexis NE, Jaspers I, Kesimer M. 2018. E-cigarette use causes a unique innate immune response in the lung, involving increased neutrophilic activation and altered mucin secretion. Am J Respir Crit Care Med. 197(4):492–501.

Subica AM, Guerrero E, Wu LT, Aitaoto N, Iwamoto D, Moss HB. 2020. Electronic cigarette use and associated risk factors in u.S.-dwelling pacific islander young adults. Subst Use Misuse. 55(10):1702–1708.

Tomar SL, Fox CH, Connolly GN. 2015. Electronic cigarettes: The tobacco industry’s latest threat to oral health? J Am Dent Assoc. 146(9):651–653.

Vallone DM, Cuccia AF, Briggs J, Xiao H, Schillo BA, Hair EC. 2020. Electronic cigarette and juul use among adolescents and young adults. JAMA Pediatr.

Walker N, Parag V, Wong SF, Youdan B, Broughton B, Bullen C, Beaglehole R. 2020. Use of e-cigarettes and smoked tobacco in youth aged 14-15 years in new zealand: Findings from repeated cross-sectional studies (2014-19). Lancet Public Health. 5(4):e204–e212.

Wamamili B, Wallace-Bell M, Richardson A, Grace RC, Coope P. 2020. Electronic cigarette use among university students aged 18-24 years in new zealand: Results of a 2018 national cross-sectional survey. BMJ Open. 10(6):e035093.

